# Identifying drug targets for schizophrenia through gene prioritization

**DOI:** 10.1101/2024.05.15.24307423

**Authors:** Julia Kraft, Alice Braun, Swapnil Awasthi, Georgia Panagiotaropoulou, Marijn Schipper, Nathaniel Bell, Danielle Posthuma, Antonio F. Pardiñas, Schizophrenia Working Group of the Psychiatric Genomics Consortium, Stephan Ripke, Karl Heilbron

**Author notes:** Equal contribution (Julia Kraft, Alice Braun). Equal contribution (Stephan Ripke, Karl Heilbron).

## Abstract

**Background:** Schizophrenia genome-wide association studies (GWASes) have identified >250 significant loci and prioritized >100 disease-related genes. However, gene prioritization efforts have mostly been restricted to locus-based methods that ignore information from the rest of the genome.

**Methods:** To more accurately characterize genes involved in schizophrenia etiology, we applied a combination of highly-predictive tools to a published GWAS of 67,390 schizophrenia cases and 94,015 controls. We combined both locus-based methods (fine-mapped coding variants, distance to GWAS signals) and genome-wide methods (PoPS, MAGMA, ultra-rare coding variant burden tests). To validate our findings, we compared them with previous prioritization efforts, known neurodevelopmental genes, and results from the PsyOPS tool.

**Results:** We prioritized 62 schizophrenia genes, 41 of which were also highlighted by our validation methods. In addition to *DRD2*, the principal target of antipsychotics, we prioritized 9 genes that are targeted by approved or investigational drugs. These included drugs targeting glutamatergic receptors (*GRIN2A* and *GRM3*), calcium channels (*CACNA1C* and *CACNB2*), and GABA_B_ receptor (*GABBR2*). These also included genes in loci that are shared with an addiction GWAS (*e.g. PDE4B* and *VRK2*).

**Conclusions:** We curated a high-quality list of 62 genes that likely play a role in the development of schizophrenia. Developing or repurposing drugs that target these genes may lead to a new generation of schizophrenia therapies. Rodent models of addiction more closely resemble the human disorder than rodent models of schizophrenia. As such, genes prioritized for both disorders could be explored in rodent addiction models, potentially facilitating drug development.

## Introduction

Schizophrenia is a highly-heritable and heterogeneous disorder characterized by positive symptoms (*e.g.* delusions and hallucinations), negative symptoms (*e.g.* blunted affect), and cognitive impairment^1^. Schizophrenia patients are often also diagnosed with neurodevelopmental disorders^1,2^ (*e.g.* intellectual disability and autism spectrum disorder) and other psychiatric conditions^3,4^ (*e.g.* substance use disorder [SUD] and depression). Antipsychotic medications antagonizing the dopamine receptor D2 are currently the first-line treatment for schizophrenia. However, approximately 34% of patients are considered treatment-resistant^5^, and especially cognitive deficits and negative symptoms often persist^6,7^. These unmet clinical needs, as well as the high burden of antipsychotic side effects^8,9^, clearly underline the necessity for pharmacotherapies with novel mechanisms of action.

Only 6.2% of psychiatric drug programs that enter Phase I trials are ultimately approved—well below the average success rate of 9.6% across all medical areas^10^—and investment in psychiatric drug development programs have decreased in recent years^11^. This low success rate likely reflects the complex nature of mental disorders, limited knowledge of disease mechanisms, and sparsity of validated animal models. Given that 63% of drugs approved by the FDA from 2013–2022 were supported by human genetic evidence^12^, pursuing targets that are genetically-linked to disease may lead to increased success rates^13,14^. A major source of this human genetic evidence comes from genome-wide association studies (GWASes)^13,14^. For instance, schizophrenia GWASes^15–17^ have identified a robust association near *DRD2,* which encodes the dopamine receptor D2. It is estimated that only 1.9% of genetically-supported drug targets for psychiatric disorders have been clinically explored^18^, suggesting that follow up of other schizophrenia GWAS findings may eventually lead to the design of new medicines.

The largest published schizophrenia GWAS identified 287 significant loci and prioritized 120 genes for follow up using fine-mapped credible sets^19^, summary data-based Mendelian randomization^20^ (SMR), and Hi-C interactions between enhancers and promoters^21^. However, these methods only use information within a given locus, ignoring information from other significant loci and the rest of the genome. The polygenic priority score (PoPS)^22^ is a gene prioritization tool that incorporates genome-wide information from MAGMA^23^ gene-level association tests and more than 57,000 gene-level features (*i.e.* gene expression, biological pathways, and protein-protein interactions). The original PoPS publication^22^ reported that it was possible to predict “probable causal genes” (defined using fine-mapped coding variants) with 79% precision and 39% recall simply by selecting genes that 1) were the nearest gene to a GWAS lead variant, and 2) had the top PoPS value in that same GWAS locus. This combined approach substantially increased precision compared to either individual approach, but with a moderate loss of recall (nearest gene: 46% precision and 48% recall; top PoPS value: 50% precision and 50% recall).

Here, we prioritized genes likely to play an important role in schizophrenia (SCZ) risk by combining PoPS and nearest gene results with additional high-precision prioritization methods—fine-mapped coding variants and ultra-rare coding variant burden tests^24^. We nominated 62 genes, 10 of which are targets of approved drugs (7 genes) or drugs that have been tested in clinical trials (“investigational drugs”, 3 genes). We discuss the potential for repurposing these drugs for schizophrenia and highlight an additional 3 prioritized genes that may be tractable via small molecule drugs.

## Methods and Materials

### Ethics statement

This research was conducted in accordance with the ethical standards of the institutional and national research committees. Informed consent was obtained from all participants.

Details on Institutional Review Board approvals of the individual studies included in the presented work are provided in the original publication^17^.

### GWAS summary statistics

We analyzed the publicly-available “core dataset” of GWAS summary statistics from the largest published SCZ GWAS from the Psychiatric Genomics Consortium (hereafter we will refer to this study as “PGC3”)^17^, a meta-analysis of 90 cohorts of European (EUR) and East Asian (EAS) descent including 67,390 cases and 94,015 controls (effective sample size [N_eff_] = up to 156,797). For analyses requiring data from a single ancestry, we used the EUR subset of the core dataset (76 cohorts, 53,386 cases, 77,258 controls, effective sample size [N_eff_] = up to 126,282) and the EAS-ancestry subset (14 cohorts, 14,004 cases, 16,757 controls, N_eff_ = up to 30,515).

### Reference panels

Accordingly, we used external data from the Haplotype Reference Consortium release 1.1 (HRC) to construct three linkage disequilibrium (LD) reference panels: an EUR panel (*N* ≥ 16,860), an EAS panel (*N* ≥ 538), and an EUR+EAS panel that included both EUR and EAS individuals in the same proportions as the GWAS summary statistics—80% EUR and 20% EAS (*N*_EUR_ = 2,191, N_EAS_ = 538).

### Variant quality control

We removed EUR+EAS GWAS variants with: 1) a minor allele count < 10 (minor allele frequency [MAF] < 0.0018) in the EUR+EAS reference panel (259 variants removed), 2) a reported allele frequency that differed from the reference panel frequency by > 0.1 (29 variants removed), and 3) a reported allele frequency that differed from the reference panel frequency by > 12-fold (11 variants removed). After quality control, 7,584,817 variants remained.

### Isolating independent association signals

In order to disentangle statistically-independent genetic signals in the EUR+EAS dataset, we first clumped variants using PLINK v1.9^25^ (*P* < 5×10^-8^, *r*^2^ < 0.1, window size = 3Mbp) and our EUR+EAS reference panel, expanded the boundaries of each clump by 500kb on either side, and merged overlapping boundaries. Within each resulting region, we ran COJO^26^ and removed hits with joint *P* > 5×10^-8^. If multiple independent hits in a region were found, we used COJO to isolate each signal by performing leave-one-hit-out conditional analysis. For each isolated signal, we computed credible sets (CSs) using the finemap.abf function in the coloc R package^27,28^. Finally, we defined loci as ±300kb around each credible set.

### MAGMA and PoPS

We performed gene-based association tests using MAGMA^23^ (“SNP-wise mean model”) and all variants with MAF > 1%. We analyzed the EUR- and EAS-based GWASes separately using the corresponding ancestry-specific reference panel and MAFs. We mapped variants to protein-coding genes using Genome Reference Consortium Human Build 37 (GRCh37) gene start and end positions from GENCODE v44^29^. We removed genes that had fewer than 3 variants mapped to them. For each gene, we meta-analyzed the resulting ancestry-specific MAGMA z-scores weighted by the square root of sample size^30^. Using the ancestry-specific MAGMA results as input, we performed PoPS^22^ using all 57,543 gene-based features as predictors. These features were not available for chrX so we restricted our analysis to autosomal genes. The resulting ancestry-specific PoPS values were then also meta-analyzed weighted by the square root of sample size. We only used the meta-analyzed MAGMA and PoPS values for gene prioritization.

### Gene prioritization criteria

Following the original PoPS publication, we prioritized genes that met both of the following criteria: 1) had the top PoPS value in a given locus and 2) were the nearest gene to the corresponding GWAS signal based on the posterior inclusion probability (PIP)-weighted average position of credible set variants. Under these criteria, however, it is possible that the top POPS value within a locus is relatively weak on a genome-wide scale, or that the nearest gene is nevertheless relatively distant. We therefore also required that genes have a PoPS value in the top 10% of all values genome-wide and the top MAGMA z-score in the locus. We also prioritized genes that had 1) PIP > 1% for non-synonymous credible set variants affecting the gene, or 2) false discovery rate-corrected P value (P_FDR_) < 5% in a published SCZ burden test of ultra-rare coding variants^24^. We used non-synonymous variants from the “baseline-LF 2.2.UKB model” (80,693 variants) and subsetted to those with an estimated per-variant heritability > 1×10^-7^ (removed 4,709 variants, all with estimated h^2^ < 1×10^-10^: >1,000-fold smaller)^31^. We removed loci that contained more than 8 genes since larger loci are more challenging to resolve^32^, but we have included results for these large loci in Table S3.

### Comparison with previous schizophrenia gene prioritization efforts

We compared our prioritized genes with those highlighted in the original PGC3 publication. Specifically, we extracted the “Symbol.ID” and “Prioritised” columns from Table S12. While the PGC3 study utilized the same core dataset, they restricted analysis to loci that retained genome-wide significance in the “extended GWAS”—a meta-analysis of the core dataset, 9 cohorts of African American and Latin American ancestry, and a dataset from deCODE genetics. They prioritized genes using a combination of FINEMAP, SMR, Hi-C interaction mapping, and non-synonymous or untranslated region credible set variants with PIP > 10%. The PGC3 study validated their list of prioritized genes by looking for overlap with genes expressed in brain tissue, genes with signatures of mutation intolerance in large-scale exome studies^33^, or genes linked to schizophrenia through rare genetic variation in the SCHEMA study^24^. Furthermore, they also found genetic overlaps in other neurodevelopmental conditions using sequencing studies from autism spectrum disorder^34^ and developmental disorder^35^. We incorporated a subset of this information by extracting the “ASD” and “DDD” columns from Table S12 of the PGC3 study. For full details, please refer to the original publication^17^.

### PsyOPS

We further validated our prioritized genes using the Psychiatric Omnilocus Prioritization Score (PsyOPS) tool^36^. The original PsyOPS publication^36^ found that PsyOPS achieved similar performance to PoPS in predicting causal psychiatric disease genes, but using only three predictors: probability of loss-of-function intolerance (pLI) > 0.99, brain-specific gene expression, and overlap with 1,370 known genes for neurodevelopmental disorders (autism, epilepsy, intellectual disability). PsyOPS treats the nearest gene to each GWAS hit as a proxy for the causal gene in the locus, trains leave-one-chromosome-out logistic regression models, and outputs the predicted probability that a given gene is causal. We determined a gene to be prioritized by PsyOPS if the predicted probability of being a causal gene exceeded 50%. We computed PsyOPS scores using all 257 independent schizophrenia GWAS hits.

### Drug repurposing and tractability

We determined whether our prioritized genes were targeted by approved or investigational drugs using GraphQL API queries of the Open Targets platform^37^, which in turn queries the EMBL-EBI ChEMBL database. For genes that were not targeted by approved or investigational drugs, we performed additional Open Targets API queries to extract evidence of drug tractability—the probability of identifying a drug that is able to bind and modulate a given target. We focussed on small molecule drugs, but results for other modalities can be found in Figure S1.

### Colocalization with other studies

We prioritized several genes that have also been highlighted by recent GWASes for addiction^38^ and Parkinson’s disease^39^. Using the EUR reference panel, we processed EUR-ancestry GWAS summary statistics from these studies using the same pipeline described above. We identified loci that physically overlapped with schizophrenia loci and computed the posterior probability of colocalization (H_4_) using all variants in the shared locus and the coloc.abf function in the coloc R package^27,28^.

## Results

We prioritized schizophrenia genes using the “core dataset” from the largest published schizophrenia GWAS meta-analysis^17^, “PGC3” (67,390 cases and 94,015 controls). We identified 257 independent associations with *P* < 5×10^-8^ (Table S1). Across these loci, we prioritized 62 schizophrenia genes (Figure 1, Table S2) based on their distance to the credible set, PoPS and MAGMA scores, number of genes in the locus, presence of non-synonymous variants in the credible set, and support from a published schizophrenia burden test of ultra-rare coding variants^24^ (see Methods). To validate our findings, we compared them with prioritization efforts from the PGC3 study^17^, genes linked to autism spectrum disorder^34^ (ASD) and developmental disorder^35^ (DD) via sequencing studies, and results from the PsyOPS tool (Figure 2). Across all genes in GWAS loci, prioritized genes were also DD and/or ASD genes (Fisher’s exact test P = 6.7×10^-14^, odds ratio [OR] = 67) or PsyOPS genes (Fisher’s exact test P = 3.9×10^-6^, OR = 14) significantly more often than expected due to chance.

**Figure 1.**
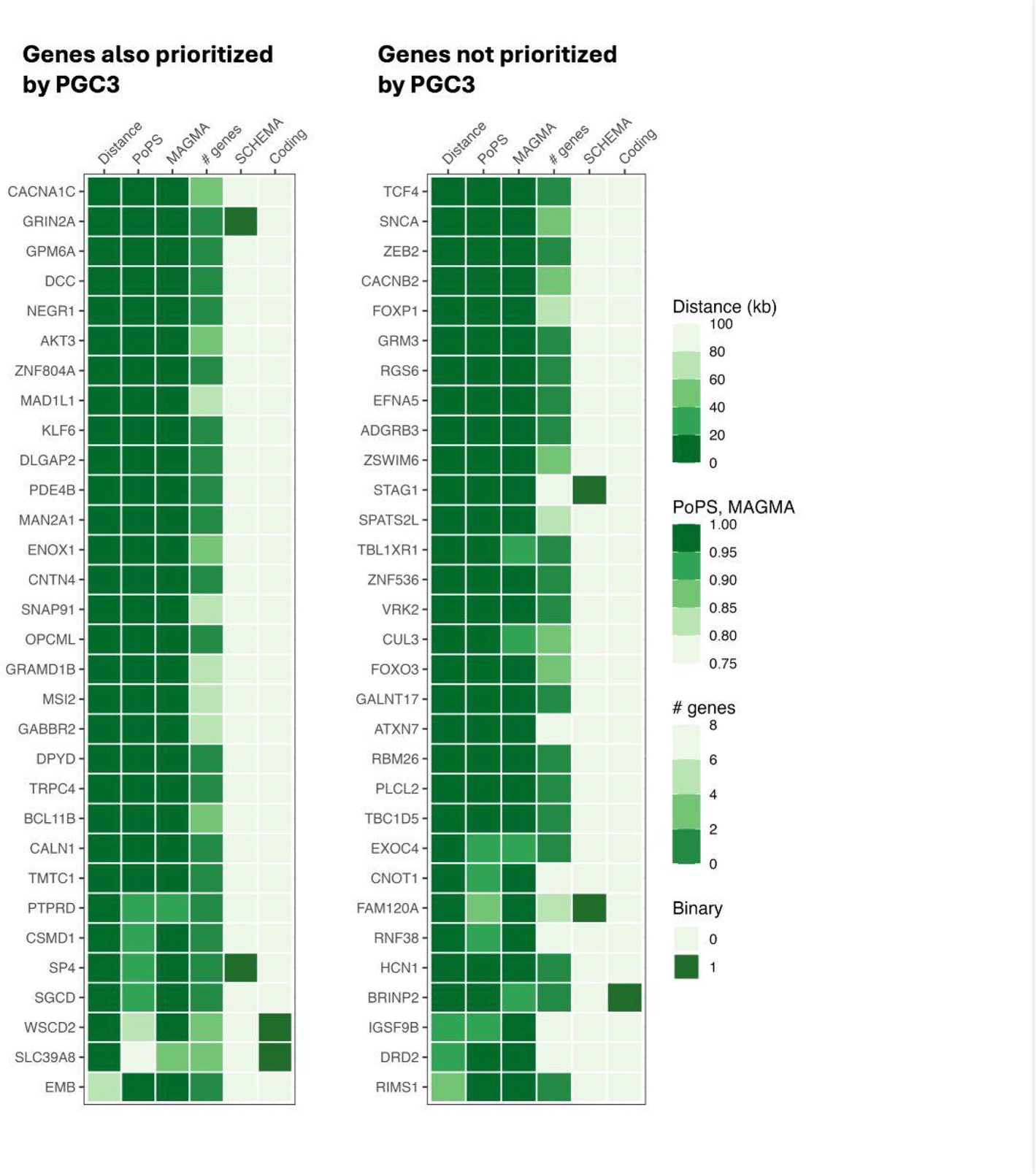
Heatmap. An overview of the evidence supporting each prioritized gene, separated based on whether they were (left panel) or were not (right panel) previously prioritized in the PGC3 study^17^. Distance: distance in kilobases between gene and credible set. PoPS: PoPS percentile where 0 represents the smallest genome-wide value and 1 represents the largest. MAGMA: MAGMA z-score percentile. # genes: number of genes in the locus. SCHEMA: a binary indicator of whether ultra-rare coding variant burden in a given gene was also significantly associated (P_FDR_ < 5%) with schizophrenia in a study from the Schizophrenia Exome Sequencing Meta-analysis (SCHEMA) consortium^24^. Coding: a binary indicator of whether the credible set contained non-synonymous variants with a summed posterior inclusion probability >1%. Genes are sorted first by distance, then by PoPS percentile.

**Figure 2.**
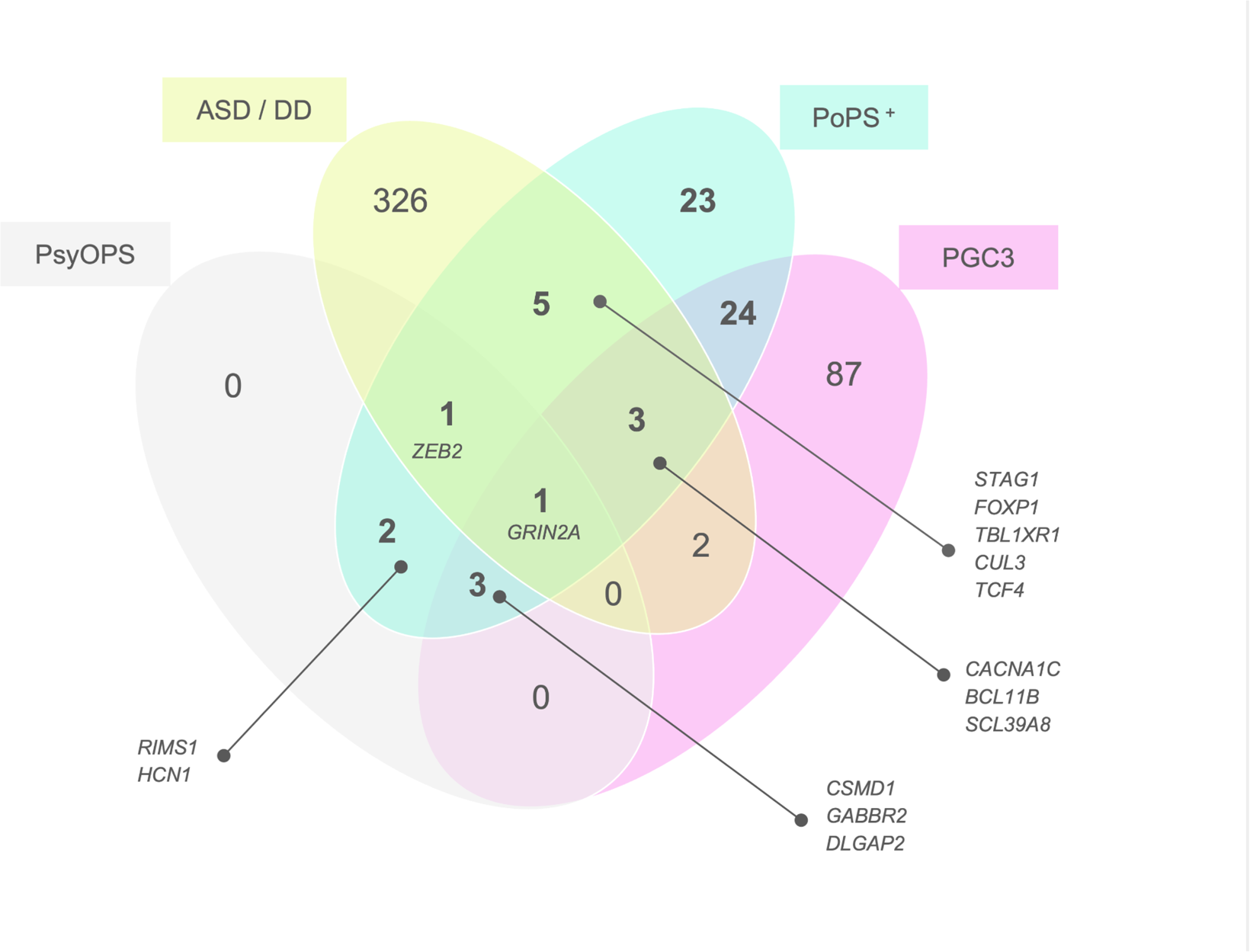
Venn diagram. Venn diagram showing the overlap between the number of genes identified by the present analysis (PoPS^+^), rare-variant studies of autism spectrum disorder (ASD) and/or developmental disorder (DD), the Psychiatric Omnilocus Prioritization Score (PsyOPS), and prior gene prioritization efforts (PGC3). Gene symbols are displayed for a subset of intersecting regions.

### Overlap with previous schizophrenia gene prioritization efforts

Of our 62 prioritized genes, 31 (50%) were also prioritized in the PGC3 study (“overlapping genes”) and several sources of evidence suggest that these genes are likely to play a role in schizophrenia risk. Ultra-rare coding variant burden in two overlapping genes (*GRIN2A* and *SP4*) was significantly associated (P_FDR_ < 5%) with schizophrenia in the SCHEMA study^24^. Similarly, four overlapping genes (*GRIN2A, CACNA1C*, *BCL11B*, and *SLC39A8*) were also identified by rare variant exome sequencing studies of DD^35^ and/or ASD^34^ (see Figure 2). Furthermore, the lead schizophrenia variant in the *SLC39A8* locus is a non-synonymous variant (PIP = 99%) that has been investigated in detail elsewhere^40^. *WSCD2* was also prioritized due to a non-synonymous variant in the credible set (PIP = 53%). Four overlapping genes (*GRIN2A*, *DLGAP2*, *GABBR2*, and *CSMD1*) were nominated by PsyOPS (see Methods). Notably, *CSMD1* is known to inhibit the complement cascade, has reduced expression in first-episode psychosis patients^41^, and knockout mice have exhibited behaviors resembling schizophrenia negative symptoms^42^.

### Genes that were not nominated by previous schizophrenia gene prioritization efforts

Of our 62 prioritized genes, 31 (50%) were not prioritized in the PGC3 study (“non-overlapping genes”). However, a similar proportion of these non-overlapping genes were supported by the same evidence sources as presented above (9/31 for overlapping genes vs. 10/31 for non-overlapping genes). Two non-overlapping genes (*STAG1* and *FAM120A*) were significantly associated with ultra-rare coding variant burden in the SCHEMA study^24^. Five non-overlapping genes (*FOXP1*, *TBL1XR1*, *ZEB2*, *CUL3*, and *TCF4*) were also identified by rare variant exome sequencing studies of DD^35^ and/or ASD^34^. Note that *TCF4* was not prioritized in the PGC3 study because they only investigated regions containing three independent genetic associations or fewer and there were four associations near *TCF4*. We prioritized *BRINP2* due to a non-synonymous variant in the credible set (r^2^ with lead variant = 97%, PIP = 2.5%), but was not prioritized in the PGC3 study which required PIP > 10%. Three non-overlapping genes (*ZEB2*, *HCN1*, and *RIMS1*) were nominated by PsyOPS (see Methods). Perhaps most importantly, our analysis uniquely highlighted the dopamine receptor gene *DRD2*, which is targeted by most approved antipsychotic medications^43^ (Figure 3A).

**Figure 3.**
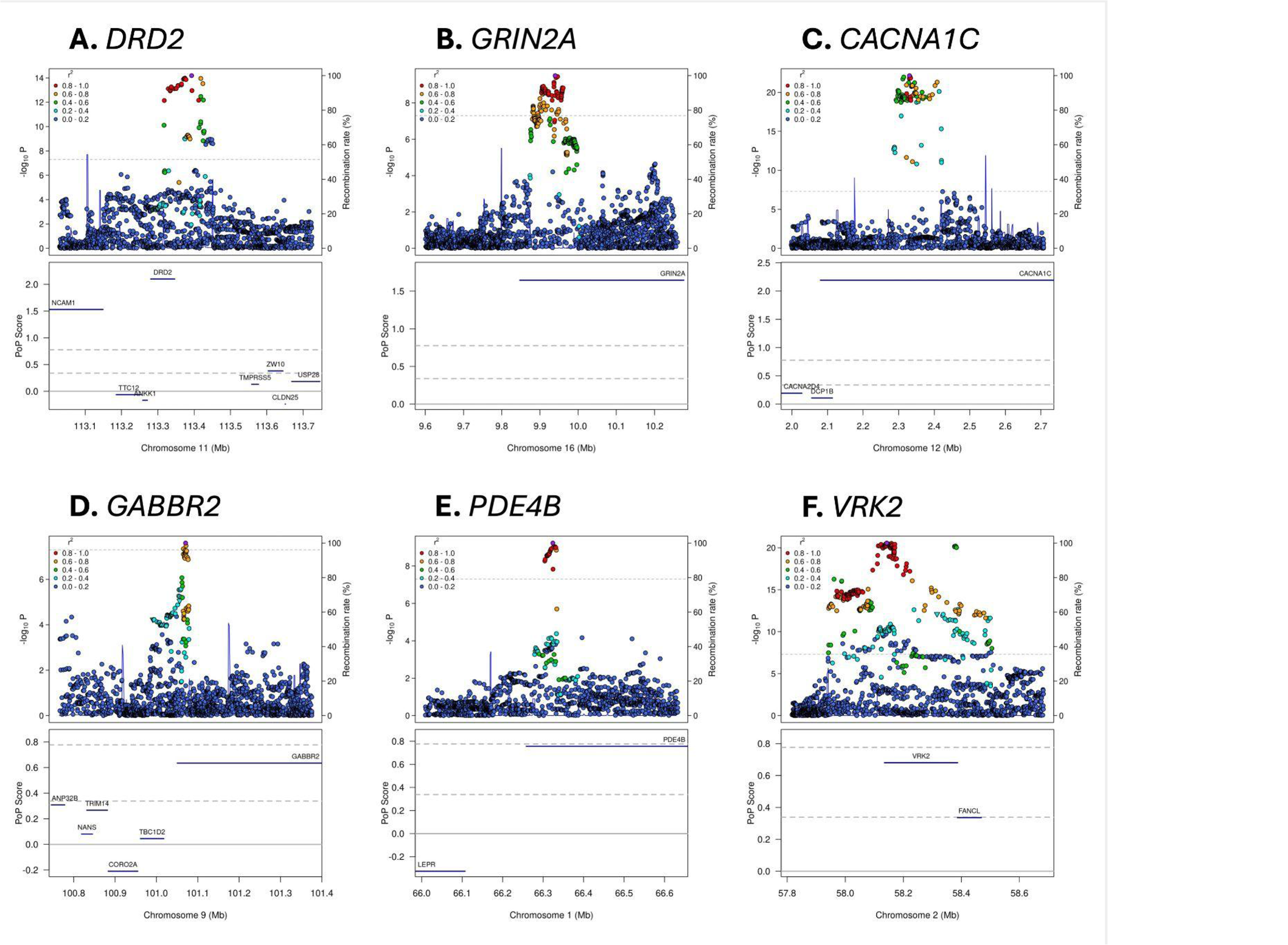
Variant-level associations and PoPS results for selected loci. The prioritized genes in plots **A-E** are targets of approved drugs; the prioritized genes in plots **E-F** are in loci shared by an addiction GWAS^38^. The upper portion of each sub-plot is a LocusZoom plot. Each point represents a different genetic variant, the x-axis represents physical position on the listed chromosome, the left y-axis represents –log_10_-transformed P value, the right y-axis represents the recombination rate, colour represents linkage disequilibrium with the lead variant in the locus (as shown in the legend), and the horizontal dashed line represents the genome-wide significance P value threshold of 5×10^-8^. The lower portion of each figure is a PoPS plot. Genes are denoted as blue bars spanning from their transcription start site to their transcription stop site using the same x-axis as the LocusZoom plot, the y-axis represents the raw PoPS score, the dashed horizontal grey lines represent the top 10% and 1% of PoPS scores genome-wide, and the solid horizontal grey line represents a PoPS score of 0.

### Drug repurposing and tractability

In addition to *DRD2*, we prioritized 9 genes that are targeted by approved (6 genes) or investigational drugs (3 genes, Table S4). Of these, 6 were also prioritized in the PGC3 study (*GRIN2A*, *CACNA1C*, *PDE4B*, *GABBR2*, *AKT3*, and *DPYD*) and 3 (*CACNB2*, *GRM3*, and *SNCA*) were uniquely prioritized in our analysis (Table 1, see Discussion). Our list of prioritized genes also included 3 genes (*HCN1*, *VRK2*, *TRPC4*) that belong to known druggable protein families^44^ and are reported to bind to at least one high-quality ligand^37^, suggesting potential as small molecule drug targets (Figure S1).

**Table 1.**
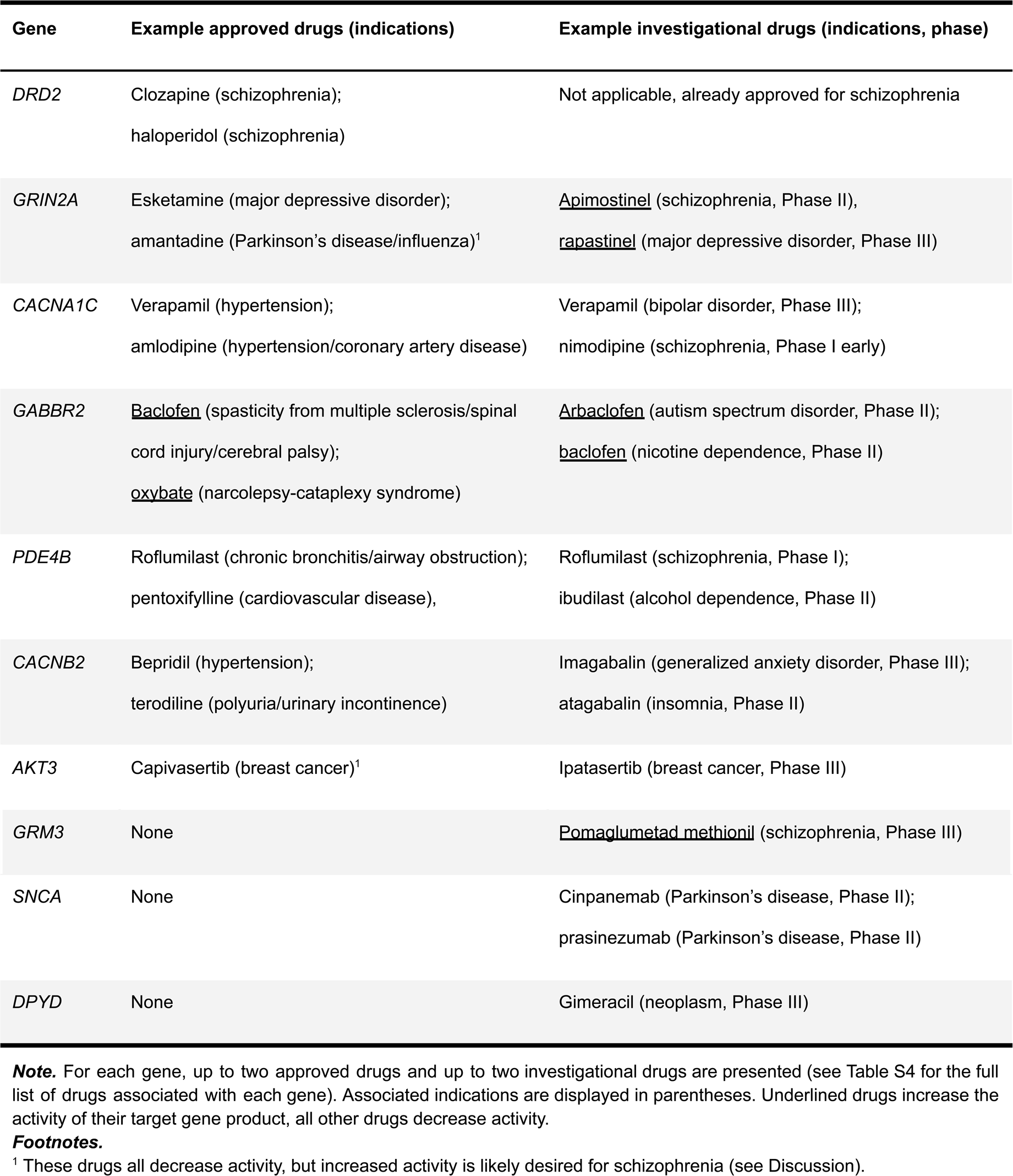
Prioritized genes targeted by approved and investigational drugs

| Gene | Example approved drugs (indications) | Example investigational drugs (indications, phase) |
| --- | --- | --- |
| *DRD2* | Clozapine (schizophrenia);<br>haloperidol (schizophrenia) | Not applicable, already approved for schizophrenia |
| *GRIN2A* | Esketamine (major depressive disorder);<br>amantadine (Parkinson's disease/influenza)<sup>1</sup> | <u>Apimostinel</u> (schizophrenia, Phase II),<br><u>rapastinel</u> (major depressive disorder, Phase III) |
| *CACNA1C* | Verapamil (hypertension);<br>amlodipine (hypertension/coronary artery disease) | Verapamil (bipolar disorder, Phase III);<br>nimodipine (schizophrenia, Phase I early) |
| *GABBR2* | <u>Baclofen</u> (spasticity from multiple sclerosis/spinal<br>cord injury/cerebral palsy);<br><u>oxybate</u> (narcolepsy-cataplexy syndrome) | <u>Arbaclofen</u> (autism spectrum disorder, Phase II);<br><u>baclofen</u> (nicotine dependence, Phase II) |
| *PDE4B* | Roflumilast (chronic bronchitis/airway obstruction);<br>pentoxifylline (cardiovascular disease), | Roflumilast (schizophrenia, Phase I);<br>ibudilast (alcohol dependence, Phase II) |
| *CACNB2* | Bepridil (hypertension);<br>terodiline (polyuria/urinary incontinence) | Imagabalin (generalized anxiety disorder, Phase III);<br>atagabalin (insomnia, Phase II) |
| *AKT3* | Capivasertib (breast cancer)<sup>1</sup> | Ipatasertib (breast cancer, Phase III) |
| *GRM3* | None | <u>Pomaglumetad methionil</u> (schizophrenia, Phase III) |
| *SNCA* | None | Cinpanemab (Parkinson's disease, Phase II);<br>prasinezumab (Parkinson's disease, Phase II) |
| *DPYD* | None | Gimeracil (neoplasm, Phase III) |

**Note.** For each gene, up to two approved drugs and up to two investigational drugs are presented (see Table S4 for the full list of drugs associated with each gene). Associated indications are displayed in parentheses. Underlined drugs increase the activity of their target gene product, all other drugs decrease activity.
<sup>1</sup> These drugs all decrease activity, but increased activity is likely desired for schizophrenia (see Discussion).

## Discussion

We prioritized 62 genes near 257 independent GWAS signals. Of these genes, 41 (66%) were also supported by evidence (Figure 2) from the PGC3 study (31 genes), DD/ASD sequencing studies (10 genes), and PsyOPS (7 genes). We prioritized *DRD2* (Figure 3A)^43^, 9 other genes targeted by approved drugs (6 genes) or drugs that have been tested in clinical trials (3 genes), and 3 other genes that may represent tractable small molecule drug targets. Our analyses do not predict whether the effect of these drugs (*e.g.* inhibitor) aligns with the effect that would be desired for schizophrenia. Therefore, we will now discuss literature supporting the potential for these drugs to be repurposed as treatments for schizophrenia.

### Glutamate receptors: GRIN2A and GRM3

We prioritized *GRIN2A*, which encodes a subunit of the N-methyl-D-aspartate receptor (NMDA-R, Figure 3B). In addition to GWAS, there is evidence that decreased NMDA-R function increases schizophrenia risk from *GRIN2A* ultra-rare variant burden tests^24^, *GRIN2A* mouse knockout models^45^, and pharmacological antagonism of the NMDA-R^46^. This raises the possibility that increasing NMDA-R activity may provide therapeutic benefit for schizophrenia patients. A meta-analysis of 4,937 schizophrenia patients from 40 randomized controlled trials found that NMDA-R modulator augmentation (*e.g.* via glycine or glycine transporter type I inhibitors) significantly improved total, positive, and negative schizophrenia symptoms versus placebo^47^. These compounds have also been proposed as a therapeutic strategy for schizophrenia patients who are treatment-resistant or have impaired cognition^48^ . There are currently three Phase III clinical trials underway assessing the effect of iclepertin, a glycine transporter type I inhibitor, on cognitive impairment associated with schizophrenia^49^. If ultimately approved, this may become the first medication indicated to treat the cognitive symptoms of schizophrenia.

We also prioritized *GRM3*, which encodes a different glutamate receptor: metabotropic glutamate receptor 3 (mGluR_3_). Clinical trials of pomaglumetad methionil, an mGluR_2/3_ agonist, have yielded inconclusive effects on positive symptoms^50,51–54^. However, an analysis of clinical trial data suggested that specific patient subgroups may have benefited^55^ and preclinical research has suggested that a cognitive endpoint may be more appropriate^56,57^.

### Voltage-gated calcium channels: CACNA1C and CACNB2

We prioritized *CACNA1C* (Figure 3C), which encodes the alpha-1 subunit of a voltage-gated calcium channel (Ca_v_1.2). A Phase III clinical trial for bipolar disorder showed that 11 out of 13 non-responders to first-line therapy (lithium) showed a clinically-meaningful response to verapamil (a calcium channel blocker [CCB]), or verapamil + lithium^58^. The genetic correlation between schizophrenia and bipolar disorder is approximately 70%^2^ and a recent bipolar disorder GWAS also identified a significant association near *CACNA1C*^59^, suggesting that verapamil may be a promising treatment option for schizophrenia. Other CCBs may also be effective—a large cohort study (N = 10,460) found that use of dihydropyridine CCBs was associated with reduced risk of psychiatric rehospitalization^60^. CCBs may also improve certain cognitive functions^61,62^. The use of CCBs for treating schizophrenia is further supported by the fact that we prioritized *CACN2B*, an auxiliary subunit of voltage-gated calcium channels.

### Loci shared with addiction: PDE4B and VRK2

We prioritized *PDE4B*, which encodes phosphodiesterase 4B (Figure 3E). A recent GWAS of an addiction-related latent factor derived from four SUDs^38^ also found a signal near *PDE4B* and highlighted *PDE4B* as the likely causal gene. SUDs are frequently comorbid with schizophrenia^4^ and there is significant genetic correlation between schizophrenia and several SUDs^63^. While it is challenging to assess psychotic symptoms in rodents, high-quality rodent addiction models exist for a wide range of substances^64^. Indeed, several drugs that are approved to treat alcohol use disorder (*e.g.* naltrexone and acamprosate) were originally pursued based in part on success in preclinical animal models^64,65^. Administering ibudilast, a drug that inhibits *PDE4B* and other phosphodiesterases, has been shown to reduce alcohol intake by approximately 50% in rats^66^ and decrease the odds of heavy drinking by 45% in a randomized clinical trial in humans^67^. Given that both addiction and schizophrenia GWASes have suggested an important role for *PDE4B* in disease risk, PDE4B inhibitors may also benefit schizophrenia patients. A Phase I study in 15 schizophrenia patients found that roflumilast, an inhibitor of all four *PDE4* phosphodiesterases, significantly improved verbal memory, but not working memory^68^.

We prioritized *VRK2*, which encodes vaccinia-related kinase 2 (Figure 3F). While the role of *VRK2* in schizophrenia remains unclear, it is expressed in microglial cells and a mechanism involving synaptic elimination by microglial cells has been proposed^69,70^. Like *PDE4B*, the same addiction GWAS^38^ also found an association near *VRK2*. The addiction and schizophrenia signals colocalize (H_4_ = 92%), suggesting a shared causal variant. Therefore, modulating VRK2 activity might result in clinical benefit for people with SUD and/or schizophrenia. VRK2 is a member of the highly-druggable serine/threonine kinases group of enzymes^44^ and has been co-crystallised with a small molecule ligand^71^. *VRK2* modulation could be tested in rodent addiction models and, if successful, may warrant further testing in human clinical trials of SUD and SCZ patients.

Three other prioritized genes reside in loci shared with the addiction GWAS^38^: *DRD2*, *SLC39A8* (H_4_ = 100%), and *PLCL2* (H_4_ = 74%). Although our analyses did not find evidence that *SLC39A8* and *PLCL2* are easily druggable by small molecule drugs, knockdown or overexpression of these genes in rodent addiction models may nevertheless improve our understanding of the shared biology of addiction and schizophrenia.

### GABBR2

We prioritized *GABBR2*, which encodes the gamma-aminobutyric acid (GABA) type B receptor and is known to inhibit neuronal activity via downstream signaling cascades (Figure 3D). A Phase II clinical trial is currently testing whether arbaclofen, a GABA_B_ receptor agonist, can rescue ASD symptoms^72^. Both post-mortem and *in vivo* studies identified reduced GABA levels in schizophrenia patients compared to controls, and impaired gamma band oscillations—which are linked with GABAergic signaling—are associated with schizophrenia^73–77^. If proven to be a successful therapy for ASD, arbaclofen may therefore represent an interesting drug repurposing candidate for schizophrenia, particularly for symptoms and socio-cognitive deficits that are shared between the two disorders^78,79^.

### AKT3

We prioritized *AKT3*, the member of the AKT serine/threonine-protein kinase gene family with the highest brain-specific expression. Capivasertib—an inhibitor of all three AKT kinases—was recently approved by the FDA to treat a subset of breast cancer patients^80^. However, AKT inhibition can lead to adverse psychiatric side effects^81^ and *AKT3* knockout or knockdown resulted in cognitive deficits and reduced brain size in mice^82,83^. Further studies are necessary to determine whether overall or isoform-specific^84^ increases in AKT3 activity would benefit schizophrenia patients without increasing cancer risk.

### SNCA

We prioritized *SNCA*, which encodes α-synuclein (α-syn). α-syn aggregates are the pathological hallmark of Parkinson’s disease (PD) and antibodies targeting aggregated α-syn have been tested in two Phase II clinical trials for PD, although neither meet their primary endpoint^85,86^. The schizophrenia association near *SNCA* colocalizes (H_4_ = 85%) with an association from a recent European-ancestry PD GWAS^39^. The schizophrenia risk allele was associated with increased PD risk, which is in turn linked to increased α-syn production^87^. As such, interventions that decrease α-syn production may benefit both PD and schizophrenia patients.

### Limitations

The PGC3 study prioritized 89 genes that were not prioritized in our study. The majority of these (52 genes) were prioritized via SMR. We did not include SMR because it demonstrated lower precision than other methods in predicting a “gold standard” dataset of causal and non-causal trait-gene pairs^22^, consistent with recent models for systematic differences between variants highlighted by GWAS and expression studies^88^. The precision of SMR-nominated genes that failed to meet our gene prioritization criteria is likely to be lower still. The PGC3 study also prioritized 5 autosomal genes affected by non-synonymous credible set variants: *ACTR1B, CUL9, IRF3, THAP8,* and *ZNF835*. These genes resided in “large loci” (containing >8 genes), which are intrinsically harder to resolve^32^. However, these genes may warrant further attention given that coding variants have been shown to prioritize causal genes with high precision^89^. An additional 10 genes met all of our prioritization criteria, but resided in large loci. Of these, 2 were prioritized by the PGC3 study (*FURIN* and *ACE*) and 8 were not (*YWHAE*, *CACNA1I*, *CHRNA3*, *AGO3*, *KIF21B*, *PTPRF*, *SYNGAP1*, and *GATAD2B*). *CACNA1I*, *CHRNA3*, and *ACE* may be particularly interesting since they are targeted by approved drugs and may represent drug repurposing opportunities.

The original PGC3 study performed gene prioritization analyses in the “core dataset”. This excluded individuals of African (AFR) or Latin American (LAT) ancestry found in the “extended dataset”. To ensure consistency with the original PGC3 study, we also analyzed the core dataset. Furthermore, the AFR and LAT datasets only included GWAS summary statistics, not individual-level genotypes, preventing us from identifying well-matched LD reference panels—something particularly important for admixed populations^90^. Nevertheless, we stress the importance of expanding gene prioritization to include more ancestries to ensure that findings are generalizable to a broader range of people.

## Conclusion

We have curated a high-quality list of 62 genes that likely play a role in the development of schizophrenia. Developing or repurposing drugs that target these genes may lead to a new generation of schizophrenia therapies. The highest-priority candidates nominated by our work and previous clinical trials are NMDA-R modulator augmentation (*GRIN2A*) and brain-penetrant calcium channel blockers (*CACNA1C* and *CACNB2*). We prioritized genes that likely also play a role in SUD, including *PDE4B* and *VRK2*. Drugs that modulate the activity of these genes should be tested in high-quality rodent models of addiction and, if shown to be safe and effective, should be considered for human clinical trials for SUD and/or schizophrenia. As new drug modalities continue to be invented and refined, more genes will become druggable. We hope that our list of prioritized genes will ultimately facilitate the development of new medicines for people living with schizophrenia.

## Data Availability

All data produced in the study are contained in the manuscript, supplementary files or available upon reasonable request to the authors. All data used in the manuscript are available online at the following websites.

https://www.ebi.ac.uk/chembl/

https://ega-archive.org/datasets/EGAD00001002729

https://www.gencodegenes.org/human/release_44.html

https://platform-docs.opentargets.org/

https://pgc.unc.edu/for-researchers/data-access-committee/data-access-portal/

https://pgc.unc.edu/for-researchers/download-results/

## Acknowledgements

We thank SURF (www.surf.nl) for the support in using the Snellius National Supercomputer.

JK and SR were supported by the German Center for Mental Health (DZPG). AB, JK, AFP, and SR were supported by the European Union’s Horizon program (101057454, “PsychSTRATA”). AB and SR were supported by The German Research Foundation (402170461, grant “TRR265”). DP and MS were supported by The Netherlands Organization for Scientific Research (NWO Gravitation: BRAINSCAPES: A Roadmap from Neurogenetics to Neurobiology - Grant No. 024.004.012). DP was supported by The European Research Council (Advanced Grant No ERC-2018-AdG GWAS2FUNC 834057). AFP, NB, and DP were supported by the European Union’s Horizon program (964874, “REALMENT”). AFP was supported by an Academy of Medical Sciences “Springboard” award (SBF005\1083). KH was supported by a Humboldt Research Fellowship from the Alexander von Humboldt Foundation. GP, SA, DP, SR, and the research reported in this publication were supported by the National Institute Of Mental Health of the National Institutes of Health under Award Number R01MH124873. The content is the responsibility of the authors and does not necessarily represent the official views of the National Institutes of Health.

*Members of the Schizophrenia Working Group of the Psychiatric Genomics Consortium* Vassily Trubetskoy, Antonio F Pardiñas, Georgia Panagiotaropoulou, Swapnil Awasthi, Tim B Bigdeli, Charlotte A Dennison, Lynsey S Hall, Max Lam, Oleksandr Frei, Alexander L Richards, Jakob Grove, Zhiqiang Li, Mark Adams, Ingrid Agartz, Elizabeth G Atkinson, Esben Agerbo, Mariam Al Eissa, Margot Albus, Madeline Alexander, Behrooz Z Alizadeha, Köksal Alptekin, Thomas D Als, Farooq Amin, Volker Arolt, Manuel Arrojo, Lavinia Athanasiu, Maria Helena Azevedo, Silviu A Bacanu, Nicholas J Bass, Martin Begemann, Richard A Belliveau, Judit Bene, Beben Benyamin, Sarah E Bergen, Giuseppe Blasi, Julio Bobes, Stefano Bonassi, Alice Braun, Rodrigo Affonseca Bressan, Evelyn J Bromet, Richard Bruggeman, Peter F Buckley, Randy L Buckner, Jonas Bybjerg-Grauholm, Wiepke Cahn, Murray J Cairns, Monica E Calkins, Vaughan J Carr, David Castle, Stanley V Catts, Kimberley D Chambert, Raymond CK Chan, Boris Chaumette, Wei Cheng, Eric FC Cheung, Siow Ann Chong, David Cohen, Angèle Consoli, Quirino Cordeiro, Javier Costas, Charles Curtis, Michael Davidson, Kenneth L Davis, Lieuwe de Haan, Franziska Degenhardt, Lynn E DeLisi, Ditte Demontis, Faith Dickerson, Dimitris Dikeos, Timothy Dinan, Srdjan Djurovic, Jubao Duan, Giuseppe Ducci, Johan G Eriksson, Lourdes Fañanás, Stephen V Faraone, Alessia Fiorentino, Andreas Forstner, Josef Frank, Nelson B Freimer, Menachem Fromer, Alessandra Frustaci, Ary Gadelha, Giulio Genovese, Elliot S Gershon, Marianna Giannitelli, Ina Giegling, Paola Giusti-Rodríguez, Stephanie Godard, Jacqueline I Goldstein, Javier González Peñas, Ana González-Pinto, Srihari Gopal, Jacob Gratten, Michael F Green, Tiffany A Greenwood, Olivier Guillin, Sinan Gülöksüz, Raquel E Gur, Ruben C Gur, Blanca Gutiérrez, Eric Hahn, Hakon Hakonarson, Vahram Haroutunian, Annette M Hartmann, Carol Harvey, Caroline Hayward, Frans A Henskens, Stefan Herms, Per Hoffmann, Daniel P Howrigan, Masashi Ikeda, Conrad Iyegbe, Inge Joa, Antonio Julià, Anna K Kähler, Tony Kam-Thong, Yoichiro Kamatani, Sena Karachanak-Yankova, Oussama Kebir, Matthew C Keller, Brian J Kelly, Andrey Khrunin, Sung-Wan Kim, Janis Klovins, Nikolay Kondratiev, Bettina Konte, Julia Kraft, Michiaki Kubo, Vaidutis Kučinskas, Zita Ausrele Kučinskiene, Agung Kusumawardhani, Hana Kuzelova-Ptackova, Stefano Landi, Laura C Lazzeroni, Phil H Lee, Sophie E Legge, Douglas S Lehrer, Rebecca Lencer, Bernard Lerer, Miaoxin Li, Jeffrey Lieberman, Gregory A Light, Svetlana Limborska, Chih-Min Liu, Jouko Lönnqvist, Carmel M Loughland, Jan Lubinski, Jurjen J Luykx, Amy Lynham, Milan Macek Jr, Andrew Mackinnon, Patrik KE Magnusson, Brion S Maher, Wolfgang Maier, Dolores Malaspina, Jacques Mallet, Stephen R Marder, Sara Marsal, Alicia R Martin, Lourdes Martorell, Manuel Mattheisen, Robert W McCarley, Colm McDonald, John J McGrath, Helena Medeiros, Sandra Meier, Bela Melegh, Ingrid Melle, Raquelle I Mesholam-Gately, Andres Metspalu, Patricia T Michie, Lili Milani, Vihra Milanova, Marina Mitjans, Espen Molden, Esther Molina, María Dolores Molto, Valeria Mondelli, Carmen Moreno, Christopher P Morley, Gerard Muntané, Kieran C Murphy, Inez Myin-Germeys, Igor Nenadić, Gerald Nestadt, Liene Nikitina-Zake, Cristiano Noto, Keith H Nuechterlein, Niamh Louise O’Brien, F Anthony O’Neill, Sang-Yun Oh, Ann Olincy, Vanessa Kiyomi Ota, Christos Pantelis, George N Papadimitriou, Mara Parellada, Tiina Paunio, Renata Pellegrino, Sathish Periyasamy, Diana O Perkins, Bruno Pfuhlmann, Olli Pietiläinen, Jonathan Pimm, David Porteous, John Powell, Diego Quattrone, Digby Quested, Allen D Radant, Antonio Rampino, Mark H Rapaport, Anna Rautanen, Abraham Reichenberg, Cheryl Roe, Joshua L Roffman, Julian Roth,

Matthias Rothermundt, Bart PF Rutten, Safaa Saker-Delye, Veikko Salomaa, Julio Sanjuan, Marcos Leite Santoro, Adam Savitz, Ulrich Schall, Rodney J Scott, Larry J Seidman, Sally Isabel Sharp, Jianxin Shi, Larry J Siever, Kang Sim, Nora Skarabis, Petr Slominsky, Hon-Cheong So, Janet L Sobell, Erik Söderman, Helen J Stain, Nils Eiel Steen, Agnes A. Steixner-Kumar, Elisabeth Stögmann, William S Stone, Richard E Straub, Fabian Streit, Eric Strengman, T Scott Stroup, Mythily Subramaniam, Catherine A Sugar, Jaana Suvisaari, Dragan M Svrakic, Neal R Swerdlow, Jin P Szatkiewicz, Thi Minh Tam Ta, Atsushi Takahashi, Chikashi Terao, Florence Thibaut, Draga Toncheva, Paul A Tooney, Silvia Torretta, Sarah Tosato, Gian Battista Tura, Bruce I Turetsky, Alp Üçok, Arne Vaaler, Therese van Amelsvoort, Ruud van Winkel, Juha Veijola, John Waddington, Henrik Walter, Anna Waterreus, Bradley T Webb, Mark Weiser, Nigel M Williams, Stephanie H Witt, Brandon K Wormley, Jing Qin Wu, Zhida Xu, Robert Yolken, Clement C Zai, Wei Zhou, Feng Zhu, Fritz Zimprich, Eşref Cem Atbaşoğlu, Muhammad Ayub, Alessandro Bertolino, Donald W Black, Nicholas J Bray, Gerome Breen, Nancy G Buccola, William F Byerley, Wei J Chen, C Robert Cloninger, Benedicto Crespo-Facorro, Gary Donohoe, Robert Freedman, Cherrie Galletly, Massimo Gennarelli, David M Hougaard, Hai-Gwo Hwu, Assen V Jablensky, Steven A McCarroll, Jennifer L Moran, Ole Mors, Preben B Mortensen, Bertram Müller-Myhsok, Amanda L Neil, Merete Nordentoft, Michele T Pato, Tracey L Petryshen, Ann E Pulver, Thomas G Schulze, Jeremy M Silverman, Jordan W Smoller, Eli A Stahl, Debby W Tsuang, Elisabet Vilella, Shi-Heng Wang, Shuhua Xu, Rolf Adolfsson, Celso Arango, Bernhard T Baune, Sintia Iole Belangero, Anders D Børglum, David Braff, Elvira Bramon, Joseph D Buxbaum, Dominique Campion, Jorge A Cervilla, Sven Cichon, David A Collier, Aiden Corvin, Marta Di Forti, Enrico Domenici, Hannelore Ehrenreich, Valentina Escott-Price, Tõnu Esko, Ayman H Fanous, Anna Gareeva, Micha Gawlik, Pablo V Gejman, Michael Gill, Stephen J Glatt, Vera Golimbet, Kyung Sue Hong, Christina M Hultman, Steven E Hyman, Nakao Iwata, Erik G Jönsson, René S Kahn, James L Kennedy, Elza Khusnutdinova, George Kirov, James A Knowles, Marie-Odile Krebs, Claudine Laurent-Levinson, Jimmy Lee, Todd Lencz, Douglas F Levinson, Qingqin S Li, Jianjun Liu, Anil K Malhotra, Dheeraj Malhotra, Andrew McIntosh, Andrew McQuillin, Paulo R Menezes, Vera A Morgan, Derek W Morris, Bryan J Mowry, Robin M Murray, Vishwajit Nimgaonkar, Markus M Nöthen, Roel A Ophoff, Sara A Paciga, Aarno Palotie, Carlos N Pato, Shengying Qin, Marcella Rietschel, Brien P Riley, Margarita Rivera, Dan Rujescu, Meram C Saka, Alan R Sanders, Sibylle G Schwab, Alessandro Serretti, Pak C Sham, Yongyong Shi, David St Clair, Ming T Tsuang, Jim van Os, Marquis P Vawter, Daniel R Weinberger, Thomas Werge, Dieter B Wildenauer, Xin Yu, Weihua Yue, Peter A Holmans, Panos Roussos, Evangelos Vassos, Danielle Posthuma, Ole A Andreassen, Kenneth S Kendler, Michael J Owen, Naomi R Wray, Mark J Daly, Hailiang Huang, Benjamin M Neale, Patrick F Sullivan, Stephan Ripke, James TR Walters, Michael C O’Donovan

## Data availability statement

ChEMBL Database: https://www.ebi.ac.uk/chembl/

HRC reference release 1.1: https://ega-archive.org/datasets/EGAD00001002729

Gencode release 44: https://www.gencodegenes.org/human/release_44.html

OpenTargets platform: https://platform-docs.opentargets.org/

The PGC3 GWAS core dataset is available through the PGC data access portal: https://pgc.unc.edu/for-researchers/data-access-committee/data-access-portal/

Summary statistics of the PGC3 GWAS are freely available for download: https://pgc.unc.edu/for-researchers/download-results/

## Code availability statement

Custom code used in the presented study is stored at https://github.com/kheilbron/cojo_pipe and https://github.com/kheilbron/brett

Additional software and code:

COJO: https://yanglab.westlake.edu.cn/software/gcta/#COJO coloc: https://github.com/chr1swallace/coloc

MAGMA: https://cncr.nl/research/magma/ PLINK 1.9: https://www.cog-genomics.org/plink/ PoPS: https://github.com/FinucaneLab/pops PsyOPS: https://github.com/Wainberg/PsyOPS

## References

1. Owen, M. J., Sawa, A. & Mortensen, P. B. Schizophrenia. Lancet 388, 86–97 (2016).

2. Cross-Disorder Group of the Psychiatric Genomics Consortium. Electronic address: & Cross-Disorder Group of the Psychiatric Genomics Consortium. Genomic Relationships, Novel Loci, and Pleiotropic Mechanisms across Eight Psychiatric Disorders. Cell 179, 1469–1482.e11 (2019).

3. Lu, C. et al. Large-scale real-world data analysis identifies comorbidity patterns in schizophrenia. Transl. Psychiatry 12, 154 (2022).

4. Hunt, G. E., Large, M. M., Cleary, M., Lai, H. M. X. & Saunders, J. B. Prevalence of comorbid substance use in schizophrenia spectrum disorders in community and clinical settings, 1990-2017: Systematic review and meta-analysis. Drug Alcohol Depend. 191, 234–258 (2018).

5. Potkin, S. G. et al. The neurobiology of treatment-resistant schizophrenia: paths to antipsychotic resistance and a roadmap for future research. npj Schizophrenia 6, 1–10 (2020).

6. Green, M. F., Kern, R. S., Braff, D. L. & Mintz, J. Neurocognitive deficits and functional outcome in schizophrenia: are we measuring the ‘right stuff’? Schizophr. Bull. 26, 119–136 (2000).

7. Cerveri, G., Gesi, C. & Mencacci, C. Pharmacological treatment of negative symptoms in schizophrenia: update and proposal of a clinical algorithm. Neuropsychiatr. Dis. Treat. 15, 1525–1535 (2019).

8. Chow, R. T. S. et al. An umbrella review of adverse effects associated with antipsychotic medications: the need for complementary study designs. Neurosci. Biobehav. Rev. 155, (2023).

9. Iversen, T. S. J. et al. Side effect burden of antipsychotic drugs in real life - Impact of gender and polypharmacy. Prog. Neuropsychopharmacol. Biol. Psychiatry 82, (2018).

10. Mullard, A. Parsing clinical success rates. Nat. Rev. Drug Discov. 15, 447–447 (2016).

11. Munro, J. & Dowden, H. Trends in neuroscience dealmaking. Biopharma Dealmakers (2018) doi:10.1038/d43747-020-00598-z.

12. Rusina, P. V. et al. Genetic support for FDA-approved drugs over the past decade. Nat. Rev. Drug Discov. 22, 864 (2023).

13. King, E. A., Davis, J. W. & Degner, J. F. Are drug targets with genetic support twice as likely to be approved? Revised estimates of the impact of genetic support for drug mechanisms on the probability of drug approval. PLoS Genet. 15, (2019).

14. Nelson, M. R. et al. The support of human genetic evidence for approved drug indications. Nat. Genet. 47, 856–860 (2015).

15. Schizophrenia Working Group of the Psychiatric Genomics Consortium et al. Biological Insights From 108 Schizophrenia-Associated Genetic Loci. Nature 511, 421 (2014).

16. Pardiñas, A. F. et al. Common schizophrenia alleles are enriched in mutation-intolerant genes and in regions under strong background selection. Nat. Genet. 50, (2018).

17. Trubetskoy, V. et al. Mapping genomic loci implicates genes and synaptic biology in schizophrenia. Nature 604, 502–508 (2022).

18. Minikel, E. V., Painter, J. L., Dong, C. C. & Nelson, M. R. Refining the impact of genetic evidence on clinical success. medRxiv 2023.06.23.23291765 (2023) doi:10.1101/2023.06.23.23291765.

19. Benner, C. et al. FINEMAP: efficient variable selection using summary data from genome-wide association studies. Bioinformatics 32, 1493–1501 (2016).

20. Zhu, Z. et al. Integration of summary data from GWAS and eQTL studies predicts complex trait gene targets. Nat. Genet. 48, (2016).

21. Belton, J. M. et al. Hi-C: a comprehensive technique to capture the conformation of genomes. Methods 58, (2012).

22. Weeks, E. M. et al. Leveraging polygenic enrichments of gene features to predict genes underlying complex traits and diseases. Nat. Genet. 55, 1267–1276 (2023).

23. de Leeuw, C. A., Mooij, J. M., Heskes, T. & Posthuma, D. MAGMA: generalized gene-set analysis of GWAS data. PLoS Comput. Biol. 11, e1004219 (2015).

24. Singh, T. et al. Rare coding variants in ten genes confer substantial risk for schizophrenia. Nature 604, 509–516 (2022).

25. Chang, C. C. et al. Second-generation PLINK: rising to the challenge of larger and richer datasets. Gigascience 4, 7 (2015).

26. Yang, J. et al. Conditional and joint multiple-SNP analysis of GWAS summary statistics identifies additional variants influencing complex traits. Nat. Genet. 44, 369–75, S1–3 (2012).

27. Giambartolomei, C. et al. Bayesian test for colocalisation between pairs of genetic association studies using summary statistics. PLoS Genet. 10, e1004383 (2014).

28. Wallace, C. Eliciting priors and relaxing the single causal variant assumption in colocalisation analyses. PLoS Genet. 16, e1008720 (2020).

29. Frankish, A. et al. GENCODE reference annotation for the human and mouse genomes. Nucleic Acids Res. 47, D766–D773 (2019).

30. Willer, C. J., Li, Y. & Abecasis, G. R. METAL: fast and efficient meta-analysis of genomewide association scans. Bioinformatics 26, 2190–2191 (2010).

31. Weissbrod, O. et al. Functionally informed fine-mapping and polygenic localization of complex trait heritability. Nat. Genet. 52, 1355–1363 (2020).

32. Wang, Q. S. & Huang, H. Methods for statistical fine-mapping and their applications to auto-immune diseases. Semin. Immunopathol. 44, 101–113 (2022).

33. Lek, M. et al. Analysis of protein-coding genetic variation in 60,706 humans. Nature 536, 285–291 (2016).

34. Satterstrom, F. K. et al. Large-Scale Exome Sequencing Study Implicates Both Developmental and Functional Changes in the Neurobiology of Autism. Cell 180, 568–584.e23 (2020).

35. Kaplanis, J. et al. Evidence for 28 genetic disorders discovered by combining healthcare and research data. Nature 586, 757–762 (2020).

36. Wainberg, M., Merico, D., Keller, M. C., Fauman, E. B. & Tripathy, S. J. Predicting causal genes from psychiatric genome-wide association studies using high-level etiological knowledge. Mol. Psychiatry 27, 3095–3106 (2022).

37. Koscielny, G. et al. Open Targets: a platform for therapeutic target identification and validation. Nucleic Acids Res. 45, D985–D994 (2017).

38. Hatoum, A. S. et al. Multivariate genome-wide association meta-analysis of over 1 million subjects identifies loci underlying multiple substance use disorders. Nat Ment Health 1, 210–223 (2023).

39. Nalls, M. A. et al. Identification of novel risk loci, causal insights, and heritable risk for Parkinson’s disease: a meta-analysis of genome-wide association studies. Lancet Neurol. 18, 1091–1102 (2019).

40. Mealer, R. G. et al. The schizophrenia-associated variant in SLC39A8 alters protein glycosylation in the mouse brain. Mol. Psychiatry 27, 1405–1415 (2022).

41. Hatzimanolis, A. et al. Deregulation of complement components C4A and CSMD1 peripheral expression in first-episode psychosis and links to cognitive ability. Eur. Arch. Psychiatry Clin. Neurosci. 272, 1219–1228 (2022).

42. Steen, V. M. et al. Neuropsychological deficits in mice depleted of the schizophrenia susceptibility gene CSMD1. PLoS One 8, e79501 (2013).

43. Wang, S. et al. Structure of the D2 dopamine receptor bound to the atypical antipsychotic drug risperidone. Nature 555, 269–273 (2018).

44. Finan, C. et al. The druggable genome and support for target identification and validation in drug development. Sci. Transl. Med. 9, (2017).

45. Farsi, Z. et al. Brain-region-specific changes in neurons and glia and dysregulation of dopamine signaling in Grin2a mutant mice. Neuron 111, 3378–3396.e9 (2023).

46. Wasserthal, S. et al. Effects of NMDA-receptor blockade by ketamine on mentalizing and its neural correlates in humans: a randomized control trial. Sci. Rep. 13, 17184 (2023).

47. Goh, K. K., Wu, T.-H., Chen, C.-H. & Lu, M.-L. Efficacy of -methyl--aspartate receptor modulator augmentation in schizophrenia: A meta-analysis of randomised, placebo-controlled trials. J. Psychopharmacol. 35, 236–252 (2021).

48. de Bartolomeis, A., Vellucci, L., Austin, M. C., De Simone, G. & Barone, A. Rational and Translational Implications of D-Amino Acids for Treatment-Resistant Schizophrenia: From Neurobiology to the Clinics. Biomolecules 12, (2022).

49. Rosenbrock, H., Desch, M. & Wunderlich, G. Development of the novel GlyT1 inhibitor, iclepertin (BI 425809), for the treatment of cognitive impairment associated with schizophrenia. Eur. Arch. Psychiatry Clin. Neurosci. 273, 1557–1566 (2023).

50. Dogra, S., Putnam, J. & Conn, P. J. Metabotropic glutamate receptor 3 as a potential therapeutic target for psychiatric and neurological disorders. Pharmacol. Biochem. Behav. 221, 173493 (2022).

51. Downing, A. M. et al. A Double-Blind, Placebo-Controlled Comparator Study of LY2140023 monohydrate in patients with schizophrenia. BMC Psychiatry 14, 351 (2014).

52. Adams, D. H., Zhang, L., Millen, B. A., Kinon, B. J. & Gomez, J.-C. Pomaglumetad Methionil (LY2140023 Monohydrate) and Aripiprazole in Patients with Schizophrenia: A Phase 3, Multicenter, Double-Blind Comparison. Schizophr. Res. Treatment 2014, 758212 (2014).

53. Kinon, B. J. et al. A multicenter, inpatient, phase 2, double-blind, placebo-controlled dose-ranging study of LY2140023 monohydrate in patients with DSM-IV schizophrenia. J. Clin. Psychopharmacol. 31, 349–355 (2011).

54. Patil, S. T. et al. Activation of mGlu2/3 receptors as a new approach to treat schizophrenia: a randomized Phase 2 clinical trial. Nat. Med. 13, 1102–1107 (2007).

55. Kinon, B. J., Millen, B. A., Zhang, L. & McKinzie, D. L. Exploratory analysis for a targeted patient population responsive to the metabotropic glutamate 2/3 receptor agonist pomaglumetad methionil in schizophrenia. Biol. Psychiatry 78, (2015).

56. Dogra, S. et al. Activating mGlu Metabotropic Glutamate Receptors Rescues Schizophrenia-like Cognitive Deficits Through Metaplastic Adaptations Within the Hippocampus. Biol. Psychiatry 90, 385–398 (2021).

57. Jin, L. E. et al. mGluR2 versus mGluR3 Metabotropic Glutamate Receptors in Primate Dorsolateral Prefrontal Cortex: Postsynaptic mGluR3 Strengthen Working Memory Networks. Cereb. Cortex 28, 974–987 (2018).

58. Mallinger, A. G. et al. Verapamil augmentation of lithium treatment improves outcome in mania unresponsive to lithium alone: preliminary findings and a discussion of therapeutic mechanisms. Bipolar Disord. 10, 856–866 (2008).

59. Mullins, N. et al. Genome-wide association study of more than 40,000 bipolar disorder cases provides new insights into the underlying biology. Nat. Genet. 53, 817–829 (2021).

60. Lintunen, J., Lähteenvuo, M., Tiihonen, J., Tanskanen, A. & Taipale, H. Adenosine modulators and calcium channel blockers as add-on treatment for schizophrenia. NPJ Schizophr. 7, 1 (2021).

61. Vahdani, B. et al. Adjunctive Raloxifene and Isradipine Improve Cognitive Functioning in Patients With Schizophrenia: A Pilot Study. J. Clin. Psychopharmacol. 40, (2020).

62. Zink, C. F. et al. Nimodipine improves cortical efficiency during working memory in healthy subjects. Transl. Psychiatry 10, 372 (2020).

63. Greco, L. A., Reay, W. R., Dayas, C. V. & Cairns, M. J. Pairwise genetic meta-analyses between schizophrenia and substance dependence phenotypes reveals novel association signals with pharmacological significance. Transl. Psychiatry 12, 403 (2022).

64. Spanagel, R. Animal models of addiction. Dialogues Clin. Neurosci. 19, 247–258 (2017).

65. Antonelli, M., Sestito, L., Tarli, C. & Addolorato, G. Perspectives on the pharmacological management of alcohol use disorder: Are the approved medications effective? Eur. J. Intern. Med. 103, 13–22 (2022).

66. Bell, R. L. et al. Ibudilast reduces alcohol drinking in multiple animal models of alcohol dependence. Addict. Biol. 20, 38–42 (2015).

67. Grodin, E. N. et al. Ibudilast, a neuroimmune modulator, reduces heavy drinking and alcohol cue-elicited neural activation: a randomized trial. Transl. Psychiatry 11, 355 (2021).

68. Gilleen, J. et al. An experimental medicine study of the phosphodiesterase-4 inhibitor, roflumilast, on working memory-related brain activity and episodic memory in schizophrenia patients. Psychopharmacology 238, 1279–1289 (2021).

69. Tesli, M. et al. VRK2 gene expression in schizophrenia, bipolar disorder and healthy controls. Br. J. Psychiatry 209, 114–120 (2016).

70. Lee, J. et al. Vaccinia-related kinase 2 plays a critical role in microglia-mediated synapse elimination during neurodevelopment. Glia 67, 1667–1679 (2019).

71. Ochoa, D. et al. The next-generation Open Targets Platform: reimagined, redesigned, rebuilt. Nucleic Acids Res. 51, D1353–D1359 (2023).

72. Parellada, M. et al. A Phase II randomized, double-blind, placebo-controlled Study of the efficacy, safety, and tolerability of arbaclofen administered for the treatment of social function in children and adolescents with autism spectrum disorders: Study protocol for AIMS-2-TRIALS-CT1. Front. Psychiatry 12, 701729 (2021).

73. Grent-’t-Jong, T. et al. Resting-state gamma-band power alterations in schizophrenia reveal E/I-balance abnormalities across illness-stages. Elife 7, (2018).

74. Fatemi, S. H., Folsom, T. D. & Thuras, P. D. Deficits in GABA(B) receptor system in schizophrenia and mood disorders: a postmortem study. Schizophr. Res. 128, 37–43 (2011).

75. Orhan, F. et al. CSF GABA is reduced in first-episode psychosis and associates to symptom severity. Mol. Psychiatry 23, 1244–1250 (2018).

76. Rowland, L. M. et al. In vivo measurements of glutamate, GABA, and NAAG in schizophrenia. Schizophr. Bull. 39, 1096–1104 (2013).

77. Chiu, P. W. et al. In vivo gamma-aminobutyric acid and glutamate levels in people with first-episode schizophrenia: A proton magnetic resonance spectroscopy study. Schizophr. Res. 193, 295–303 (2018).

78. De Crescenzo, F. et al. Autistic Symptoms in Schizophrenia Spectrum Disorders: A Systematic Review and Meta-Analysis. Front. Psychiatry 10, 78 (2019).

79. Oliver, L. D. et al. Social Cognitive Performance in Schizophrenia Spectrum Disorders Compared With Autism Spectrum Disorder: A Systematic Review, Meta-analysis, and Meta-regression. JAMA Psychiatry 78, (2021).

80. Mullard, A. FDA approves first-in-class AKT inhibitor. Nat. Rev. Drug Discov. 23, 9 (2024).

81. Tsimberidou, A.-M. et al. AKT inhibition in the central nervous system induces signaling defects resulting in psychiatric symptomatology. Cell Biosci. 12, 56 (2022).

82. Bergeron, Y. et al. Genetic Deletion of Akt3 Induces an Endophenotype Reminiscent of Psychiatric Manifestations in Mice. Front. Mol. Neurosci. 10, 102 (2017).

83. Howell, K. R., Floyd, K. & Law, A. J. PKBγ/AKT3 loss-of-function causes learning and memory deficits and deregulation of AKT/mTORC2 signaling: Relevance for schizophrenia. PLoS One 12, e0175993 (2017).

84. Bhattacharya, A. et al. Isoform-level transcriptome-wide association uncovers genetic risk mechanisms for neuropsychiatric disorders in the human brain. Nat. Genet. 55, 2117–2128 (2023).

85. Lang, A. E. et al. Trial of Cinpanemab in Early Parkinson’s Disease. N. Engl. J. Med. 387, 408–420 (2022).

86. Pagano, G. et al. Trial of Prasinezumab in Early-Stage Parkinson’s Disease. N. Engl. J. Med. 387, 421–432 (2022).

87. Soldner, F. et al. Parkinson-associated risk variant in distal enhancer of α-synuclein modulates target gene expression. Nature 533, 95–99 (2016).

88. Mostafavi, H., Spence, J. P., Naqvi, S. & Pritchard, J. K. Systematic differences in discovery of genetic effects on gene expression and complex traits. Nat. Genet. 55, 1866–1875 (2023).

89. Mountjoy, E. et al. An open approach to systematically prioritize causal variants and genes at all published human GWAS trait-associated loci. Nat. Genet. 53, 1527–1533 (2021).

90. Kanai, M. et al. Meta-analysis fine-mapping is often miscalibrated at single-variant resolution. Cell Genom 2, (2022).

