## Supplementary Figures for "Identifying drug targets for schizophrenia through gene prioritization"

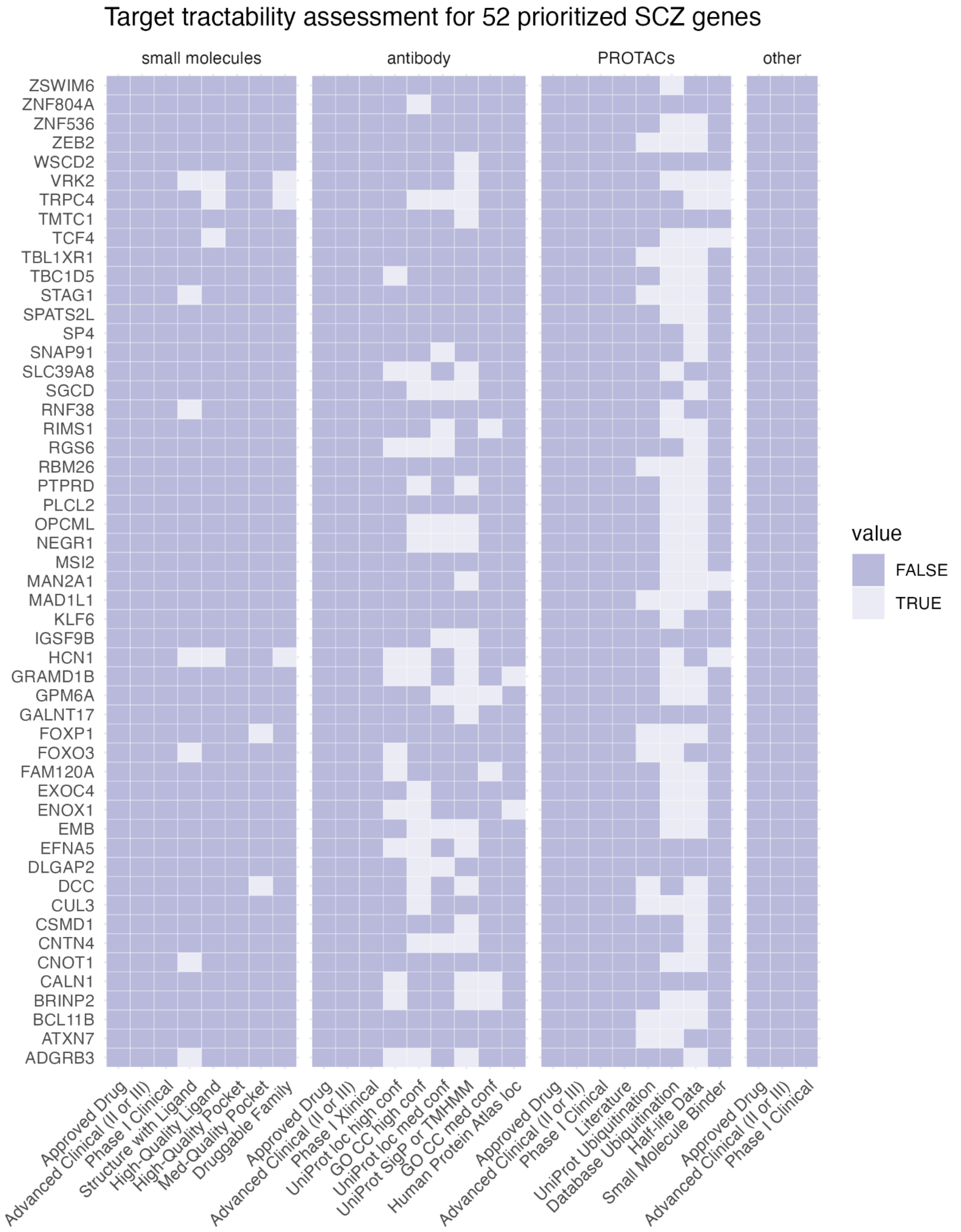


**Supplementary Figure 1**. Tractability of 52 prioritized schizophrenia (SCZ) genes (excluding 10 genes targeted by approved or investigational drugs) as novel drug targets by different modalities: small molecules, antibody, Proteolysis Targeting Chimeras (PROTACs) and other clinical modalities. Data was extracted from the Open Targets platform using GraphQL API queries (see Methods). Tractability buckets (x-axis) are presented in descending order of evidence quality within each modality.
